# Epigenetic age acceleration is associated with brain connectivity changes and the risk of major depression after trauma

**DOI:** 10.64898/2026.09.18.26363409

**Authors:** Alba Navarro-Flores, Hazel Milla, Sarah D. Linnstaedt, Xinming An, Jennifer S. Stevens, Laura Huff, Nathaniel G. Harnett, Alyssa R. Roeckner, Katelyn I. Oliver, David R. Rubinow, Karestan C. Koenen, Urs Heilbronner, Thomas G. Schulze, Kerry J. Ressler, Samuel A. McLean, Anthony S. Zannas

## Abstract

**Background:** Major depressive disorder (MDD) is associated with brain dysfunction and age-related diseases. Epigenetic clocks based on DNA methylation patterns estimate biological aging and aim to predict morbidity and mortality. Although MDD has been linked to epigenetic age acceleration (EAA) cross-sectionally, it is unknown whether EAA is associated with MDD after trauma.

**Methods:** We included 706 participants (66% non-white, 64% female) from the multi-ancestry AURORA study, enrolled at emergency departments (ED) within 72 hours after trauma, from whom three age-adjusted epigenetic clocks were measured at enrollment: PhenoAge, GrimAge, and DunedinPACE. Functional MRI and assessment for probable MDD were conducted two weeks and six months later, respectively. Logistic and linear regression models tested the association of EAA with MDD and functional neuroimaging endpoints.

**Results:** A total of 660 participants had available assessments of probable MDD six months after trauma, and from those 160 screened positively (24.2%) after trauma. EAA calculated by GrimAge (OR = 1.44, 95% CI = 1.12 – 1.85, p-adj= 0.012) and DunedinPACE (OR = 1.46, 95% CI = 1.15 – 1.85, p-adj = 0.005), but not PhenoAge (p-adj= 0.239), was significantly associated with MDD 6-months after trauma. Associations remained significant after adjusting for additional confounders, including prior trauma and disability (p= 0.008 for GrimAge, p= 0.010 for DunedinPACE). Subgroup analysis based on a median age split revealed these effects were only observed in participants 35 years of age or older. DunedinPACE was significantly associated with altered connectivity of fourteen brain network pairs after FDR correction (n = 217).

**Conclusions:** EAA is associated with a higher risk of MDD after trauma and with concomitant brain connectivity changes. Calculation of GrimAge and DunedinPACE at the time of ED presentation may help recognize patients at risk for MDD, potentially improving personalized care after trauma.

## 1. Introduction

Excessive stress and traumatic experiences are important risk factors for major depressive disorder (MDD). However, high variability in individual responses to trauma exists, which is hypothesized to result from the complex interaction between genome and environment.^1^ Physical health is another relevant determinant of depression, and the presence of physical conditions such as obesity, diabetes, stroke, inflammatory bowel disease, some forms of cancer, and other chronic inflammatory diseases increases the risk of developing MDD.^2^ Many of these conditions are age-related and frequently reported as comorbid to MDD, pointing to potentially shared pathophysiological pathways and highlighting that not only might MDD affect the pace of aging, but aging also presents a risk for MDD.^3^

Epigenetic factors are chemical modifications of DNA that can result from environmental exposures, are potentially heritable, and regulate gene expression and cellular functions.^4^ Through the lifetime of an individual, these modifications accumulate and play a role in the maintenance of the functionality of various systems and overall health.^5^ Alterations in DNA methylation (DNAm) patterns are crucial modifications that have been widely studied in humans and are known to change extensively over time, representing a hallmark of aging.^5^ Chronological age is the major risk factor for the development of cardiovascular, cerebrovascular, metabolic, neoplastic, and neurodegenerative disorders, which are leading causes of morbidity and mortality worldwide.^6, 7^ However, there is high between-subject heterogeneity in age-related disease risk.^8^ Biological aging is thought to capture this interindividual variability and to represent a more precise and personalized measure of physiological integrity and lifespan.^9^ In light of this, a new definition of aging was recently proposed as the “process resulting from accumulation of consequences of life, including molecular and cellular damage,” which contribute to functional decline, disease, and death.^10^

Biological age can be estimated using molecular aging clocks, with epigenetic clocks based on DNAm being the most established.^11^ While early epigenetic clocks were developed to predict chronological age, second generation clocks were further trained to predict morbidity and mortality outcomes. PhenoAge was developed to predict what is called “phenotypic age” by adding nine aging-relevant blood parameters to the calculation (albumin, creatinine, glucose, C-reactive protein, lymphocytic percentage and total leucocyte counts, erythrocyte and platelet distribution width, and alkaline phosphatase).^12, 13^ GrimAge was trained to estimate a value of biological age that correlates with mortality risk by utilizing specific CpG sites as surrogates that correlate with plasma levels of seven proteins (i.e., adrenomedullin, C-reactive protein, plasminogen activator inhibitor-1, etc.) and smoking pack-years.^13^ More recently, a third-generation clock – DunedinPACE or “pace of aging” – was developed to predict the rate of physiological decline.^14^ Epigenetic age acceleration (EAA) estimation based on these clocks has been associated with adverse environmental exposures and with poor cardiovascular, metabolic, and mental health outcomes.^15–17^ Therefore, these clocks are of emerging interest in psychiatry, not only to evaluate the effects of interventions or environmental factors on aging, but to analyze biological aging as a predictor of other outcomes.^18^ In that line, we previously found that EAA measured by GrimAge at the time of trauma was associated with posttraumatic stress disorder (PTSD) six months later.^19^ However, whether EAA could is associated with other neuropsychiatric outcomes following trauma exposure, like MDD, is unknown.

To improve understanding of the mechanisms linking aging and MDD, it is important to analyze the accompanying functional changes of specific brain regions and circuits using neuroimaging. A meta-analysis showed that MDD is associated with alterations in connectivity within the default mode network (DMN) and frontoparietal network (FPN), as well as increased connectivity between FPN and DMN.^20^ Moreover, functional connectivity in the temporal lobe and in limbic structures (hippocampus, parahippocampal area, agranular retrolimbic area) has been shown to mediate the effect of EAA on cognitive performance in older adults.^21^ Additional functional neural correlates of EAA in the context of trauma remain largely unknown.

In a previous analysis of a smaller subset of the AURORA, the exposure to trauma was associated with higher PTSD risk 6 months later in patients with EAA measured by GrimAge.^19^ In the present study, we evaluated the association of epigenetic aging at the time of trauma exposure and the development of MDD 6 months later. To this end, we used a larger sample from the AURORA study and evaluated for the first time in this unique cohort three next-generation epigenetic clocks (PhenoAge, GrimAge, and DunedinPACE) as predictors of MDD risk, as well as the functional brain correlates of EAA.

## 2. Methods and Materials

### Participants

The participants included in this study took part in the AURORA study.^22^ The AURORA study focused on understanding the development of adverse posttraumatic neuropsychiatric sequelae, including depression, in a total sample of 3,829 patients who consulted the emergency department (ED) up to 72 h after trauma exposure. The recruitment of the AURORA study took place between September 2017 and December 2020 in 30 EDs across the USA. ^23^ Participants signed an informed consent before participanting in the study. All study procedures were approved by the relevant central and local Institutional Review Board. A diagram of the study flow is presented in **Figure 1**.

**Figure 1.**
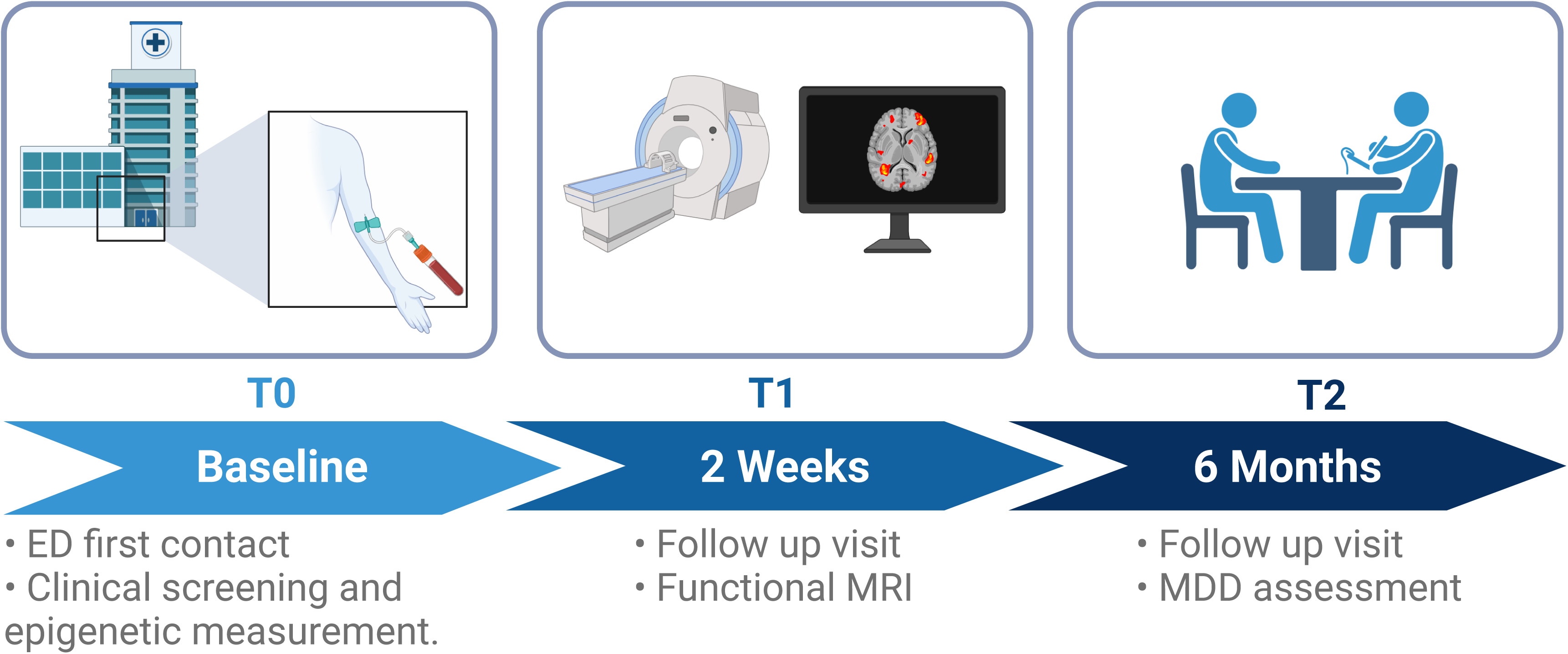
Workflow of AURORA study procedures. Participants were recruited at the emergency department after exposure to psychological trauma, and blood was collected for biomarker assessment. Neuroimaging data were collected two weeks later, and depressive symptoms were assessed six months after ED presentation. ED: emergency department; MRI: magnetic resonance imaging; MDD: major depressive disorder.

#### Inclusion criteria

Participants were adults, aged 18 to 75 years, who presented to an emergency department (ED) during the first 72 hours after exposure to a traumatic event. The included trauma exposures were motor vehicle collision, physical assault, sexual assault, fall greater than 10 feet, or mass casualty incidents. Details of the trauma types included and their sample sizes are displayed in Supplementary Table 1 (Supplement). Other exposures qualified if (1) they were reported as involving actual or threatened serious injury, sexual violence, or death, independently of it being by direct exposure, witnessing, or learning about it; or (2) under the discretion of the evaluation of the research assistant. Finally, eligible participants had an iOS or Android-compatible smartphone, internet access, and an email address for regular communication.

#### Exclusion criteria

Patients were excluded if they received general anesthesia, were taking ≥20 mg of morphine or an equivalent per day, or presented with long bone fractures, laceration with significant hemorrhage, or solid organ injury (> American Association for the Surgery of Trauma Grade 1). Those that were not alert and oriented at the time of enrollment, not fluent in written or spoken English, or presented visual or auditory impairment precluding completion of web-based neurocognitive evaluations and/or telephone follow-ups were also excluded. Finally, exclusion criteria also included those with exposures that were self-inflicted or of occupational nature, and prisoners, individuals pregnant or breastfeeding, or reporting ongoing domestic violence. The exclusion of these subgroups was done a priori based on the AURORA study protocol to reduce participant burden and heterogeneity and to focus the cohort on a more uniform traumatic-exposure phenotype, on early post-trauma enrollment and excluding factors that would complicate interpretation of post-trauma biology, especially hospitalization-related confounding.

The current study includes all AURORA participants with available epigenetic assessments at the time of ED presentation. From those, a smaller subset underwent structural and functional neuroimaging 2 weeks after the initial ED presentation.

### Phenotyping

Probable Major Depressive Disorder (abbreviated in the text as MDD) diagnosis was evaluated using the PROMIS Depression – Short Form 8b test.^24, 25^ Briefly, the scale consists of 8 items, each scored using a 1–5 response scale (none of the time, a little, some, most, and all or almost all the time).^26, 27^ Raw scores were converted to T-scores and a cutoff of 60+ (i.e., one standard deviation above the mean in the general US population) was used for dichotomizing the variable into MDD present or absent. Pre-trauma depression status was evaluated using the PROMIS-8b questionnaire, asking participants for symptoms in the 30 days prior the index traumatic event. The dichotomized values were calculated similarly as for current MDD.

Other variables used for the sensitivity analysis included measures related to trauma. The Injury Severity Score (ISS), which was calculated based on the Abbreviated Injury Scale,^28^ measured multiple injuries in the most affected body regions. The overall level of disability 30 days before the traumatic event was measured using the Sheehan Disability Scale (SDS).^29–31^ The Area Deprivation Index (ADI) was included using the ADI National Percentile Ranking 2019 as a measure of socioeconomic disadvantage.^32^ Previous traumatic experiences were measured with the total scores of the Life Events Checklist (LEC-5)^33, 34^ and 11 out of 28 items of the Childhood Trauma Questionnaire – Short form (CTQ-SF).^35^

### DNA Methylation

#### Processing of the sample

DNA methylation (DNAm) was carried as previously described.^19^ Briefly, ED blood samples, obtained in PAXgene tubes and frozen at –20°C, were batch-shipped to NIH repository on dry ice. DNA was isolated (Chemagic 360), bisulfite-converted (Zymo), and profiled (Illumina EPIC) with randomization. More details regarding the DNAm assay, quality control, and batch correction details are in Supplementary Methods (Supplement).

#### Description of the clocks used

The beta values of the processed DNAm data were used for the calculation of the epigenetic clocks using the online Horvath calculator. The values obtained were regressed against chronological age, and residuals were extracted. These standardized values were used for each clock. The three epigenetic clocks included in this study were PhenoAge (estimation of phenotypic age and risk of morbidity),^12^ GrimAge (estimation of the risk of mortality),^13^ and DunedinPACE (estimation of the pace of aging).^14^ Because blood cell composition can influence epigenetic age, models were adjusted for proportions of CD8+ T cells, CD4+ T cells, B cells, natural killer cells, granulocytes, and monocytes, estimated with a standard DNAm-based pipeline.^36^

### Genetic Ancestry Principal Components

The Infinium Global Screening Array-24 v1.0 (Illumina Inc.) was used to genotype DNA samples at the Stanley Center/Broad Institute. After quality control (Supplementary Methods – Supplement), principal component Analysis (PCA) was performed in Plink1.9. We created plink.eigenvec and plink.eigenval files after conducting PCA of our samples. Together with samples obtained from the 1000 Genomes Project (1000G), the top 10 principal components (PCs) were extracted. A decision tree was trained using the 1000G PCs, and the AURORA PCs were tested to determine genetic ancestry and the possible genetic ancestry probabilities using the R package “rpart.”

### Functional Neuroimaging

Resting-state functional MRI (rs-fMRI) data were processed as described previously,^37^ collected from 5 sites at the 2-week follow-up timepoint. Although 6-Month follow-up fMRI data are available for a smaller subset of the AURORA study, here we included the 2-week fMRI data to maximize power for detecting associations with epigenetic clock measures. Preprocessing used fMRIPrep v1.2.2 with ICA-AROMA for motion correction,^38–40^ followed by AFNI 3dTproject for detrending, filtering (0.01–0.1 Hz), and nuisance regression. Yeo 7-network ROI correlations (21 pairs, z-transformed) were used to quantify connectivity (Supplementary methods – Supplement).^41^ The coarser 7-network resolution was chosen to map trauma phenotypes to broad, interpretable functional domains rather than finer subnetworks (Yeo-17). We chose between-network correlations as our primary analysis because they capture large-scale network disintegration, a hallmark of depression and aging-related vulnerability, that is more theoretically relevant to EAA and MDD than within-network homogeneity or global segregation. ^20^

### Statistical Analysis

Statistical analyses were conducted using R version 4.3.2. We used logistic regression to test the association of chronological age-adjusted epigenetic aging measured by three clocks at the ED with a probable MDD diagnosis 6 months later. These models were adjusted by pre-trauma MDD score, self-reported sex assigned at birth, self-reported race or ethnicity, education, marital status, and DNAm-estimated blood cell proportions. Ethnicity/race was included as a covariate to account for the fact that participants of European ancestry were overrepresented for the development of the clocks, and because these variables have been associated with different biological aging trajectories. ^42^

Correction for multiple testing was done using the Bonferroni method,^43^ leading to an adjusted α (p value threshold) of 0.05/3 = 0.0167.

For the secondary analyses, we conducted the same models for the three clocks, but in patients who were 35 years old or younger and in patients older than 35 years separately (based on a median split). In sensitivity analyses, the main models were additionally adjusted for the injury severity score, level of previous disability, the area deprivation index, previous trauma (the list of threatening experiences and the childhood trauma questionnaire), and the first three genetic principal components. Two additional sensitivity analyses were conducted separately, adjusting the main models for trauma subtype (Supplementary Table 1, Supplement) and for smoking status (smoker/non-smoker). Smoking status was defined as non-smoker when patients reported not having consumed tobacco products in the previous 30 days to the index ED visit.

We further tested the association of epigenetic clocks with resting state fMRI data, using the connectivity of the 21 network pairs.^41^ All models involving neuroimaging were also adjusted by imaging site, and models for structural models were further adjusted for total intracranial volume. Correction for multiple testing was performed using the False Discovery Rate method.^44^

Odds ratios were calculated per standard deviation increase in each continuous predictor to improve comparability across variables measured on different scales. For each predictor, the logistic regression coefficient was multiplied by that variable’s standard deviation and exponentiated to obtain the OR per SD; 95% confidence intervals were derived using the coefficient standard error and the same standard deviation scaling.

## 3. Results

### 3.1. Description of the cohort

There was a total of 706 participants with epigenetic data at the time of ED presentation. The majority were non-Hispanic Black women, with a mean age of 38 and a median of 35 (which was used for the median split for the sensitivity analysis) and presented to the ED after a motor vehicle collision (80%). The three epigenetic clocks PhenoAge (Pearson’s r = 0.90, p-value < 0.0001), GrimAge (r = 0.91, p-value < 0.0001), and DunedinPACE (r =0.32, p-value < 0.0001) significantly correlated with chronological age. For the main analyses, we included those with available probable depression assessments at the 6^th^ month (n=660). Participants who developed MDD after six months, compared to those who did not, were younger, with a higher proportion of females, and presented higher scores of disability, and childhood and lifetime trauma. The description of the sample included is presented in **Table 1**.

**Table 1.**
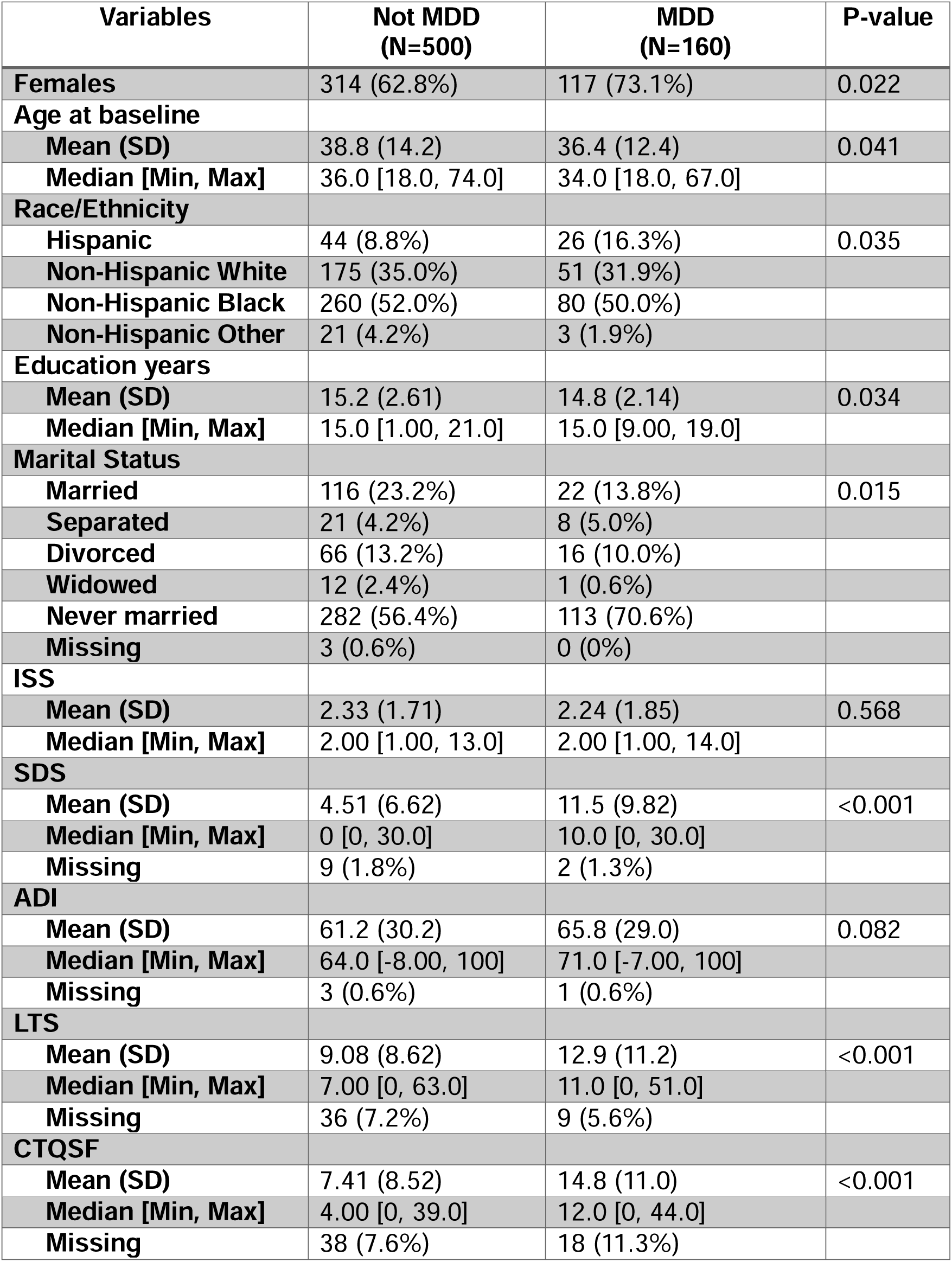
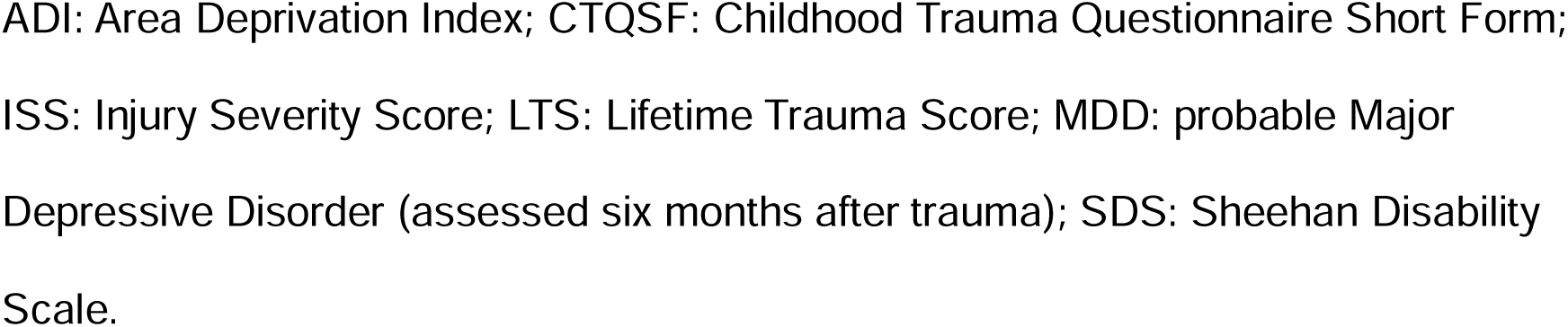
Demographic and clinical description of the participants.

| Variables | Not MDD<br>(N=500) | MDD<br>(N=160) | P-value |
| --- | --- | --- | --- |
| **Females** | 314 (62.8%) | 117 (73.1%) | 0.022 |
| **Age at baseline** |  |  |  |
| Mean (SD) | 38.8 (14.2) | 36.4 (12.4) | 0.041 |
| Median [Min, Max] | 36.0 [18.0, 74.0] | 34.0 [18.0, 67.0] |  |
| **Race/Ethnicity** |  |  |  |
| Hispanic | 44 (8.8%) | 26 (16.3%) | 0.035 |
| Non-Hispanic White | 175 (35.0%) | 51 (31.9%) |  |
| Non-Hispanic Black | 260 (52.0%) | 80 (50.0%) |  |
| Non-Hispanic Other | 21 (4.2%) | 3 (1.9%) |  |
| **Education years** |  |  |  |
| Mean (SD) | 15.2 (2.61) | 14.8 (2.14) | 0.034 |
| Median [Min, Max] | 15.0 [1.00, 21.0] | 15.0 [9.00, 19.0] |  |
| **Marital Status** |  |  |  |
| Married | 116 (23.2%) | 22 (13.8%) | 0.015 |
| Separated | 21 (4.2%) | 8 (5.0%) |  |
| Divorced | 66 (13.2%) | 16 (10.0%) |  |
| Widowed | 12 (2.4%) | 1 (0.6%) |  |
| Never married | 282 (56.4%) | 113 (70.6%) |  |
| Missing | 3 (0.6%) | 0 (0%) |  |
| **ISS** |  |  |  |
| Mean (SD) | 2.33 (1.71) | 2.24 (1.85) | 0.568 |
| Median [Min, Max] | 2.00 [1.00, 13.0] | 2.00 [1.00, 14.0] |  |
| **SDS** |  |  |  |
| Mean (SD) | 4.51 (6.62) | 11.5 (9.82) | <0.001 |
| Median [Min, Max] | 0 [0, 30.0] | 10.0 [0, 30.0] |  |
| Missing | 9 (1.8%) | 2 (1.3%) |  |
| **ADI** |  |  |  |
| Mean (SD) | 61.2 (30.2) | 65.8 (29.0) | 0.082 |
| Median [Min, Max] | 64.0 [-8.00, 100] | 71.0 [-7.00, 100] |  |
| Missing | 3 (0.6%) | 1 (0.6%) |  |
| **LTS** |  |  |  |
| Mean (SD) | 9.08 (8.62) | 12.9 (11.2) | <0.001 |
| Median [Min, Max] | 7.00 [0, 63.0] | 11.0 [0, 51.0] |  |
| Missing | 36 (7.2%) | 9 (5.6%) |  |
| **CTQSF** |  |  |  |
| Mean (SD) | 7.41 (8.52) | 14.8 (11.0) | <0.001 |
| Median [Min, Max] | 4.00 [0, 39.0] | 12.0 [0, 44.0] |  |
| Missing | 38 (7.6%) | 18 (11.3%) |  |

ADI: Area Deprivation Index; CTQSF: Childhood Trauma Questionnaire Short Form; ISS: Injury Severity Score; LTS: Lifetime Trauma Score; MDD: probable Major Depressive Disorder (assessed six months after trauma); SDS: Sheehan Disability Scale.

### 3.2. Accelerated aging measured by GrimAge and DunedinPACE at the time of trauma is associated with probable MDD six months later

We tested whether three epigenetic clocks (PhenoAge, GrimAge, and DunedinPACE) measured at ED presentation were associated with MDD six months later. Following adjustment for pre-trauma MDD score, sex assigned at birth, race or ethnicity, education, marital status, and DNAm-estimated blood cell proportions, as well as Bonferroni correction for multiple testing (adjusted α < 0.0167), GrimAge (n = 652, OR = 1.44, 95% CI = 1.12 – 1.85, β = 0.08, SE = 0.03, Z = 2.9, p-adj = 0.012) and DunedinPACE (n = 652, OR = 1.46, 95% CI = 1.15 – 1.85, β = 2.97, SE = 0.95, Z = 3.14, p-adj = 0.002), but not PhenoAge (p-adj = 0.17) at ED presentation were significantly associated with probable MDD diagnosis six months later (**Figure 2**). Sensitivity analyses showed that after further adjusting for the first three genetic principal components, previous depressive symptoms, injury severity score, level of previous disability, area deprivation index, and previous trauma, MDD was still significantly associated with both GrimAge (p=0.008) and DunedinPACE (p=0.010). To better understand these associations, participants with higher vs. lower EAA were classified using a median split for GrimAge and DunedinPACE. We found that MDD was present in 26.7% of the participants with higher EAA, compared to 21.8% of those with lower EAA measured by GrimAge (i.e., the two groups had 4.9% difference in MDD risk). When using EAA measured by DunedinPACE, 26.4% of the participants with higher scores presented MDD after six months, compared to 22.1% of their peers with lower scores (i.e., the two groups had 4.3% difference in MDD risk). These associations remained significant after sensitivity analyses for the main regression models with separate adjustments for trauma subtype and smoking status (Supplementary Table 2, Supplement).

**Figure 2.**
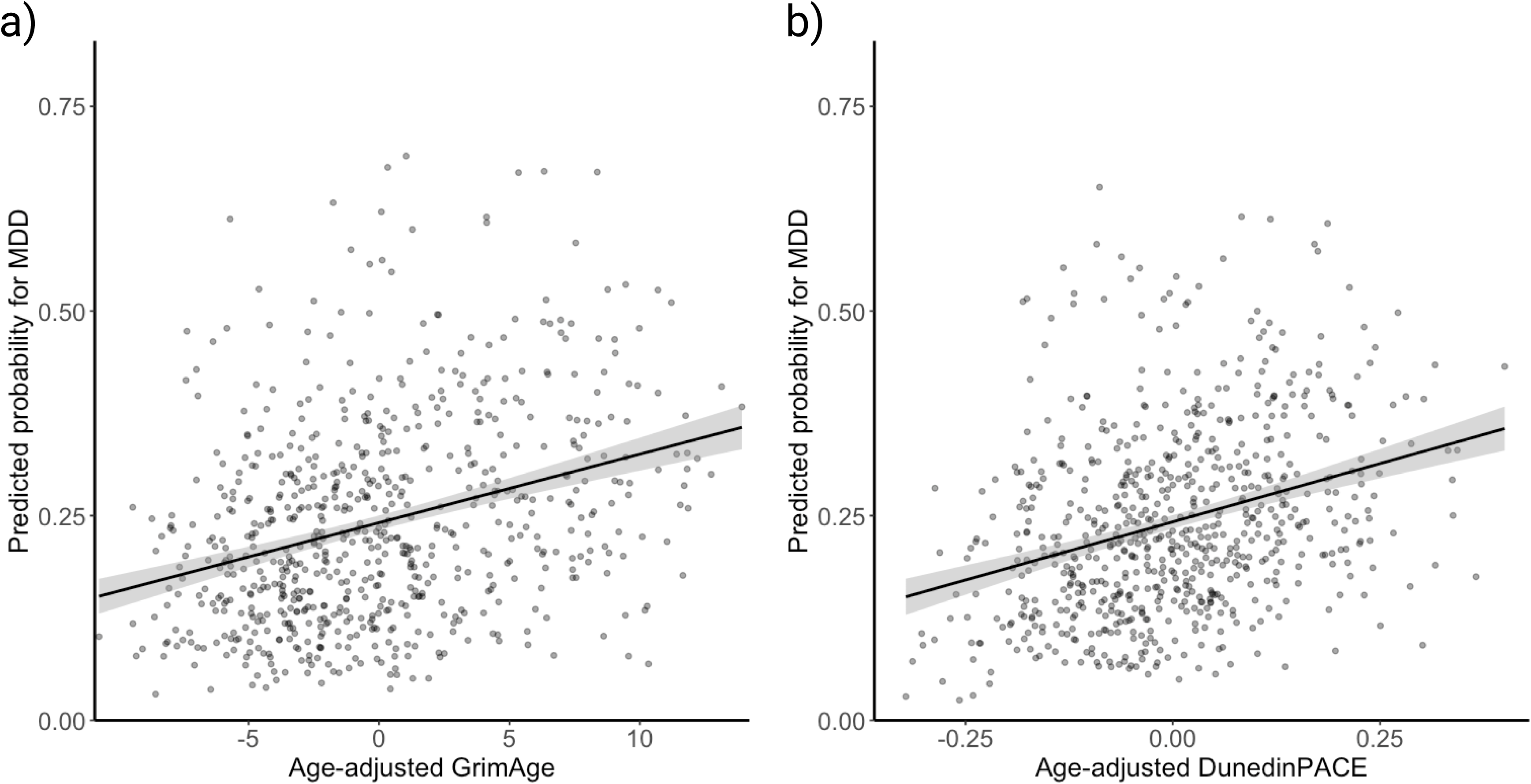
GrimAge and DunedinPACE at the time of ED presentation predict probability for 6-month MDD diagnosis 6 months later. Predicted Probabilities for MDD were calculated based on the regression models applied using age-adjusted GrimAge (a) and DunedinPACE (b) for visualization purposes. Both probabilities were predicted using logistic regression for MDD diagnosis six months after trauma, adjusted for sex assigned at birth, race/ethnicity, educational attainment, marital status, and DNAm-estimated blood cell proportions.

Secondary analyses evaluated whether these associations were modified by chronological age at the time of trauma. To maximize power, we again used a median split to create two balanced age groups. In patients older than 35 years, GrimAge (n = 326, OR = 1.5, 95% CI = 1.03 – 2.18, β = 0.08, S.E = 0.04, Z = 2.09, p = 0.036) and DunedinPACE (n = 326, OR = 1.65, 95% CI = 1.16 – 2.35, β = 3.89, S.E = 1.40, Z = 2.78, p=0.005) were significantly associated with the presence of MDD after six months. In contrast, these associations were not significant in younger individuals (n=326; p-value > 0.11). The rates of depression were similar between the younger (25.8%) and older group (22.7%), and except for educational attainment and marital status, no statistically significant differences were found in biological age acceleration rates (epigenetic clock residuals) or other confounders (Supplementary table 3, Supplement).

### 3.3. Brain connectivity associations to EAA relevant to MDD

Of the 706 patients with epigenetic data, 217 also had imaging data. Analyses involving this subset of patients showed that, after FDR correction for multiple testing, higher pace of aging measured by DunedinPACE was significantly associated with increased between-network brain connectivity of fourteen network pairs (n=217), among those the Sensorimotor-Frontoparietal, Visual-Default Mode, Ventral Attention-Frontoparietal, and the Visual-Frontoparietal network pairs had the highest coefficients (see full list in Table 2, Figure 3). Some of these rs-fMRI connectivity pairs have been associated with MDD, including the default mode and frontoparietal networks.^20^ No between-network connectivity measures were significantly associated with either PhenoAge (p-adj values ≥ 0.95) or GrimAge (p-adj values ≥ 0.06).

**Figure 3.**
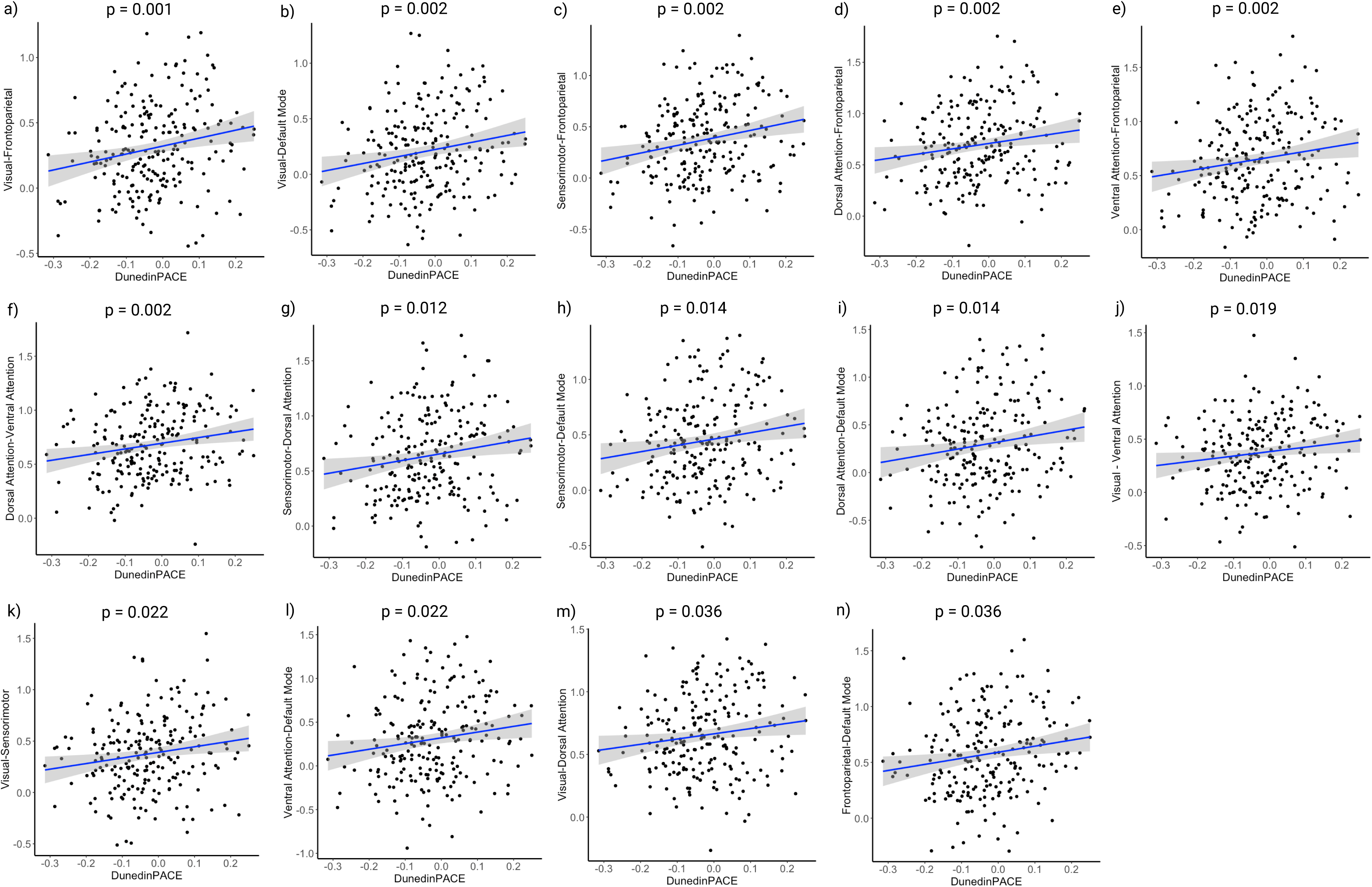
Association of the fourteen rs-MRI pairs connectivity values with DunedinPACE. The plot shows the correlation between each of the eight between-network connectivity measures measured with resting state fMRI and DunedinPACE. The significance values were calculated using linear regression models (n=217) that were adjusted for pre-trauma MDD symptoms, sex assigned at birth, race/ethnicity, education, marital status, and blood cell proportions and corrected for multiple testing with FDR.

**Table 2.** Summary of associations between functional connectivity of network pairs and DunedinPACE.

| Networks | OR (95 % CI) | β-<br>coefficient | SE | z-value | p-value |
| --- | --- | --- | --- | --- | --- |
| Visual-Frontoparietal | 1.11 (1.06 -<br>1.17) | 0.83 | 0.20 | 4.11 | 0.001 |
| Visual-Default Mode | 1.12 (1.05 -<br>1.18) | 0.86 | 0.23 | 3.75 | 0.002 |
| Sensorimotor-Frontoparietal | 1.12 (1.06 -<br>1.19) | 0.90 | 0.24 | 3.81 | 0.002 |
| Dorsal Attention-Frontoparietal | 1.10 (1.05 -<br>1.16) | 0.77 | 0.22 | 3.58 | 0.002 |
| Ventral Attention-Frontoparietal | 1.11 (1.05 -<br>1.18) | 0.85 | 0.24 | 3.53 | 0.002 |
| Dorsal Attention-Ventral<br>Attention | 1.09 (1.04 -<br>1.14) | 0.67 | 0.19 | 3.47 | 0.002 |
| Sensorimotor-Dorsal<br>Attention | 1.10 (1.03 -<br>1.17) | 0.72 | 0.25 | 2.92 | 0.012 |
| Sensorimotor-Default Mode | 1.09 (1.03 -<br>1.16) | 0.68 | 0.24 | 2.80 | 0.014 |
| Dorsal Attention-Default Mode | 1.10 (1.03 -<br>1.18) | 0.76 | 0.27 | 2.79 | 0.014 |
| **Visual-Ventral Attention** | 1.07 (1.02 – 1.13) | 0.54 | 0.21 | 2.63 | 0.019 |
| **Visual-Sensorimotor** | 1.07 (1.02 – 1.13) | 0.58 | 0.23 | 2.52 | 0.022 |
| **Ventral Attention-Default Mode** | 1.09 (1.02 – 1.17) | 0.73 | 0.29 | 2.53 | 0.022 |
| **Visual-Dorsal Attention** | 1.06 (1.01 – 1.11) | 0.45 | 0.20 | 2.27 | 0.036 |
| **Frontoparietal-Default Mode** | 1.06 (1.01 – 1.12) | 0.52 | 0.22 | 2.30 | 0.036 |

$\beta$ -coefficients represent change in z-scored network FC per unit increase in DunedinPACE residuals (all models adjusted for imaging site, sex at birth, race/ethnicity, educational attainment, marital status, cellular composition). P-values were corrected with the FDR method. CI: confidence interval, SE: standard error. Observations in each regression n = 217.

Among the EAA-associated network pairs, logistic regression (n=202) identified nominally significant associations of Visual-Default Mode Network (p = 0.004), Visual-Sensorimotor (p = 0.010), Ventral Attention-Frontoparietal (p = 0.016), Visual-Frontoparietal (p = 0.019), and the Visual-Ventral Attention between-network connectivity with probable MDD 6 months after trauma. Significance was lost after FDR correction (p-adjusted values ≥ 0.053).

## 4. Discussion

MDD is associated with accelerated biological aging measured by epigenetic clocks ^45, 46^ and brain imaging studies.^47, 48^ However, whether epigenetic clocks measured at the time of trauma are associated with the risk of developing MDD has not been explored. In this study, we used the AURORA cohort to demonstrate that accelerated aging measured by GrimAge and DunedinPACE at the time of a traumatic event is associated with MDD six months later.^22^ Moreover, we found in a subgroup analysis that these associations were only significant in patients older than 35 years, despite the MDD group being younger than the non-MDD group, suggesting that MDD risk conferred by biological aging may manifest especially when individuals are exposed to trauma at more advanced ages. Using resting state functional imaging data from a subset of participants, we discovered altered connectivity of eight network pairs in association with DunedinPACE, three of which were nominally significantly associated with MDD. These findings show that individuals with EAA are more vulnerable to developing MDD following a traumatic event and further suggest that such vulnerability may be due to EAA-associations with brain network connectivity.

GrimAge is an epigenetic clock that robustly predicts mortality.^13^ In this study, we found that EAA measured with this clock was associated with a higher risk of presenting with MDD six months after trauma. This extends our previous study from a smaller AURORA subset (n=289), which showed that accelerated GrimAge at the time of trauma was associated with PTSD outcomes six months later. ^19^ It has been estimated that PTSD and MDD co-occur in approximately 52% of patients without differences by gender, but with MDD rates being higher in patients with PTSD of military and interpersonal violence compared to those from other civilian traumatic event and those exposed to natural disasters. ^49^ This comorbidity is relevant because it is associated with worse functional outcomes, including increased suicide risk, ^50^ higher lifetime suicide attempts, comorbid anxiety disorders, and mental healthcare utilization. ^51^ Both PTSD and MDD may be linked to higher EAA due to overlapping stress-response pathways that relate to dysregulated DNA methylation patterns underlying epigenetic clocks, particularly through chronic hypothalamic-pituitary-adrenal (HPA) axis hyperactivity and inflammation that precede or are triggered by trauma exposure, ^52^ and related to the severity of psychiatric symptoms. ^53^ Together, our previous and current findings suggest that EAA may be a marker of broadly increased risk for developing neuropsychiatric disorders in response to trauma. These findings further build on relevant and emerging work in the interface of trauma, MDD, and aging. A previous study found that patients with MDD have a median of 2 years of advanced GrimAge compared to controls.^54^ The presence of higher depressive symptoms was also related to an average of 1.29 years of advanced GrimAge compared to those with low or no symptoms.^55^ Similarly, a review of nine studies identified accelerated GrimAge as a consequence of MDD,^56^ highlighting the bidirectional relationship of GrimAge acceleration and MDD diagnosis.

We further found that DunedinPACE at ED presentation was positively associated with depression six months after trauma. Other relevant risk factors related to depression that have been previously related to accelerated DunedinPACE are, for example, childhood exposure to poverty and victimization.^14^ The role of trauma was further explored in post-deployment US veterans, finding that higher DunedinPACE measured after these experiences was related to increased risk of chronic disease and mortality after follow-up.^57^ This highlights the role of epigenetic aging measured by DunedinPACE in increased vulnerability after trauma.

In our secondary analyses, we found that EAA was associated with MDD only in the group of patients older than 35 years of age. This observation may be explained by the training of next-generation epigenetic clocks in cohorts comprising, in their majority, middle-aged, older adults, and even centenarians, in whom age-related conditions are more prevalent.^12, 13^ Moreover, divergence of biological age from chronological age is more evident later in life, when the individual variability of these gaps is also more pronounced, making the predictive value of the clocks more robust in older populations. ^58^

Building on prior work suggesting increased between-network functional connectivity as a hallmark of brain aging,^59^ we found that increased connectivity in eight network pairs correlates with accelerated aging measured by DunedinPACE. Among these pairs, brain connectivity associations to the Visual-DMN, the Visual-FPN, and the Attention-FPN were further associated with 6-month probable MDD. These findings are in line with a large meta-analysis showing that MDD is associated with increased FPN-DMN between-network connectivity.^20^ In another study, the visual-DMN and visual-FPN between-network connectivity were found to be increased in a large subgroup of patients with MDD, suggesting a distinct endophenotype of depression.^60^ The increased functional connectivity involving the visual network, FPN, and DMN observed in our study may reflect heightened integration of visual sensory processing during autobiographical memory recall, a process prominently supported by the DMN. This enhanced coupling could contribute to the vivid, sensory-rich mental imagery characteristic of rumination in MDD, where DMN-FPN interactions are already known to strengthen during repetitive negative thought patterns.^61^

The association of these connectivity patterns with EAA further aligns with prior evidence linking advanced brain age to increased rumination and worry tendencies.^62^ Moreover, evidence suggests that the between-network connectivity of DMN-FPN is higher is older vs younger individuals (mid-life and late-life adults vs children, adolescents, and young-adults), which hints to the effects of aging on brain connectivity, and could provide hypothesis linking EAA to these changes. ^63^

The differential associations of epigenetic clocks with neuroimaging measures could reflect the differences in how they were developed. PhenoAge and GrimAge, which were trained on clinical biomarkers and mortality outcomes,^12, 13^ are known to be robustly associated with chronological age, yet in our analyses demonstrate weaker associations with brain connectivity. In contrast, DunedinPACE was calibrated against longitudinal multi-system decline trajectories ^14^ and is known to weaklier correlate with chronological age, yet in our analyses shows stronger brain connectivity associations. This pattern aligns with DunedinPACE’s sensitivity to current aging rates rather than accumulated damage, making it particularly relevant for capturing dynamic brain connectivity. Further studies in larger cohorts are needed to clarify the functional brain connectivity associations linking EAA and MDD.

It is important to consider some limitations for the proper interpretation of this study. The regression models of the main outcomes and the sensitivity analyses were adjusted for various relevant confounders, but the absence of a control group without trauma exposure prevented us from analyzing EAA as a mediator of MDD development in response to trauma exposure. Regarding this topic, a study using the UK Biobank cohort including more than 400,000 individuals found that those with higher biological aging were at a higher risk of presenting depression at baseline as well as developing depression in the following 8 years (for those healthy at baseline) compared to their chronologically aged peers. ^64^ Similarly, a study of more than 5000 Chinese middle-aged and older adults found not only that accelerated biological aging was associated with a higher baseline depression risk, but also that those with baseline depression later developed increased biological age compared to their healthy peers, highlighting the bidirectional relationship of biological aging and depression. ^65^ The majority of the participants were of non-white ethnicities, and the clocks analyzed had been mostly train in participants with European ancestries. To account for this, we adjusted our models by ethnicity/race, nevertheless this potential bias should be taken into consideration when interpreting these results. Other methods not used in this study for measuring biological aging such as proteomic clocks, both systemic and brain-specific, had been linked to a higher risk of incident depression in the general population.^66^

Because neuroimaging was performed 2 weeks after trauma, it is not possible to distinguish the extent to which connectivity associations to EEA are induced by the index traumatic event or were present before trauma. To evaluate connectivity changes that occur as a consequence of trauma, which could explain the development of MDD, measures before and after trauma would be needed. Additionally, a larger sample size would increase power, especially for the neuroimaging subset, in which the association between EAA and MDD was diluted, preventing us from further characterizing this association, such as through mediation analysis. Finally, longer follow-up periods and longitudinal cohort data could aid in understanding the long-term effects of trauma and disentangle the intricate interplay between trauma, MDD, and EAA.

In summary, accelerated epigenetic age at the time of trauma was associated with probable MDD six months later and was further associated with increased connectivity between brain networks known to be involved in MDD. These findings may have important implications for personalized interventions aimed at alleviating neuropsychiatric outcomes and improving the quality of life of trauma-exposed individuals.

## Data Availability

All data produced in the present study are available upon reasonable request to the authors.

## Acknowledgments

The investigators wish to thank the trauma survivors participating in the AURORA Study. Their time and effort during a challenging period of their lives make our efforts to improve recovery for future trauma survivors possible.

This project was supported by NIMH under U01MH110925, the US Army MRMC, One Mind, the Mayday Fund, and the National Institute on Aging of the National Institutes of Health under Award Number R01AG088241. This work was also supported by a NARSAD Young Investigator grant (#24135) and a Foundation of Hope for Research and Treatment of Mental Illness grant awarded to ASZ. Verily Life Sciences and Mindstrong Health provided some of the hardware and software used to perform study assessments. Bio-samples for this publication were processed by and obtained from the NIMH Repository & Genomics Resource, supported by cooperative agreement U24MH068457. Data and/or research tools used in the preparation of this manuscript were obtained from the National Institute of Mental Health (NIMH) Data Archive (NDA) under DOI: 10.15154/1528075. NDA is a collaborative informatics system created by the National Institutes of Health to provide a national resource to support and accelerate research in mental health. This manuscript reflects the views of the authors and may not reflect the opinions or views of any of the funders or of the Submitters submitting original data to NDA.

