## Supplement for "Epigenetic age acceleration is associated with brain connectivity changes and the risk of major depression after trauma"

**This is the Supplementary Material from**

Index

### **Supplementary methods**

1. **Genotyping QC**

DNA samples from AURORA participants were genotyped using the Infinium Global Screening Array-24 v1.0 (Illumina Inc.) at the Stanley Center/ Broad Institute. Genotype quality control included removing rare variants with minor allele frequency (MAF)<0.005, karyotypic abnormalities determined via B-allele frequency, cryptic relatedness with kinship coefficient (KING-robust estimates)>0.089, removing SNPs with Hardy-Weinberg equilibrium p-value<1e-8, removing SNPs with call rates <98%, and removing participants with sex chromosomes discrepant from reported sex assigned at birth.

2. **DNAm assay and QC**

After collecting blood samples at the ED in DNA PAXgene tubes (Qiagen Germantown MD, USA), those were frozen at -20C at each collection study site, for a later batch-shipping to the NIH repository preserved on dry ice (Piscataway, NJ, USA). Subsequently, DNA isolation was conducted using magnetic bead technology with the Chemagic 360 instrumentation (PerkinElmer, Waltham, MA, USA). For the calculation of DNA concentration and purity, UV/Vis on a Lunatic reader spectrophotometer were used (Unchained Labs, Pleasanton, CA, USA).

For the DNAm assay, a bisulfite conversion was done with the EZ-96 DNA Methylation Kits following the manufacturer’s instructions (Zymo Research, Irvine, CA, USA) at the University of Minnesota Genomics Center, St. Paul, MN. The quantification of DNA methylation was carried out with the Infinium Human MethylationEPIC BeadChip V1 (Illumina Inc., San Diego, CA, USA), and the samples were randomized across the arrays.

The samples were randomized samples across assay plates such in a manner that accounted for main clinical outcomes (PTSD, pain, depression), timepoint (in this study only baseline measures were used), and patient characteristics (e.g. age, sex, race).

Automated hybridization and staining were performed using a liquid handling robot (Tecan, Männedorf, Switzerland). The CHAMP package in RStudio(1) was used for quality control, filtering of low-quality probes and samples, adjustment of probe design (Infinium I and II), and batch effects.

DNAm probes with a low detection (p>0.1) or low beadcount (<3) in at least 5% of the samples were removed. Moreover, we removed cross-reactive and polymorphic probes; those containing a CpG site overlapping with SNPs, at single base extension sites, or close to short insertions or deletions; and probes located on sex chromosomes. Singular Value Decomposition analysis was performed for visual inspection of potential residual batch effects, which were removed using ComBat.(2, 3)

3. **Imaging analysis**

The preprocessing of the data (TR = 2.36 s, 230 volumes, total scan time = 9 min 5 s) was conducted using the ICA-AROMA module from the fMRIPrep v1.2.2 pipeline,(4-6) which effectively mitigates motion-related artifacts in a data-driven manner and performs as well as, or superior to, traditional scrubbing or censoring methods.

Subsequent preprocessing was conducted with the Analysis for Functional NeuroImages program (AFNI’s) 3dTproject to apply linear detrending, remove non-steady state volumes identified by FMRIPREP, perform bandpass filtering (0.01–0.1 Hz), and regress out signals from white matter, corticospinal fluid, and the global signal to reduce physiological noise.

The correlation of the mean fMRI time-course of the regions of interest (ROIs), according to the Yeo 7-Network atlas,(7) was used for the network connectivity estimation. To calculate the strength of the network-to-network functional connectivity, the Pearson correlation coefficients were calculated for 21 pairs of ROIs (each unique pair across the 7 networks) and were z-transformed before the statistical analyses.

### **Supplementary Table 1. Description of trauma subgroups**

| Categories | Type of Traumatic Event | n°  (n = 660) |
| --- | --- | --- |
| Motor Vehicle Collision | 1: Motor Vehicle Collision | 485 |
| Interpersonal Violence | 2: Physical Assault | 66 |
|  | 3: Sexual Assault | 5 |
| Other injuries | 4: Fall >= 10 feet | 11 |
|  | 5: Incident causing traumatic stress exposure to many people | 2 |
|  | 6: Non-motorized Collision | 15 |
|  | 7: Fall < 10 feet or from unknown height | 23 |
|  | 8: Poisoning | 0 |
|  | 9: Burns | 5 |
|  | 10: Animal-related | 22 |
|  | 11: Other | 26 |

### **Supplementary Table 2.** Sensitivity analysis by trauma subtype and tobacco use

| Sensitivity Analysis | OR (95 % CI) | β-coefficient | SE | z-value | p-value |
| --- | --- | --- | --- | --- | --- |
| Trauma Subtype |  |  |  |  |  |
| GrimAge | 1.41 (1.09 - 1.82) | 0.071 | 0.027 | 2.65 | 0.008 |
| PhenoAge | 1.23 (0.97 - 1.56) | 0.036 | 0.021 | 1.74 | 0.081 |
| DunedinPACE | 1.45 (1.14 - 1.83) | 2.903 | 0.954 | 3.04 | 0.002 |
| Tobacco Use |  |  |  |  |  |
| GrimAge | 1.41 (1.06 - 1.86) | 0.070 | 0.030 | 2.36 | 0.018 |
| PhenoAge | 1.24 (0.98 - 1.57) | 0.038 | 0.021 | 1.82 | 0.069 |
| DunedinPACE | 1.38 (1.08 - 1.76) | 2.549 | 0.978 | 2.61 | 0.009 |

Legend. β-coefficients represent association of epigenetic clocks with MDD (all models adjusted for previous depression symptoms, sex at birth, race/ethnicity, educational attainment, marital status, cellular composition and either trauma subtype or tobacco use). P-values were corrected with the FDR method. CI: confidence interval, SE: standard error.

### **Supplementary Table 3.** Description of the participants according to their chronological age group

|  | Older (N=331) | Younger (N=329) | P-value |
| --- | --- | --- | --- |
| Females | 221 (66.8%) | 210 (63.8%) | 0.477 |
| Age at baseline |  |  |  |
| Mean (SD) | 50.1 (8.74) | 26.4 (4.85) | <0.001 |
| Median [Min, Max] | 49.0 [36.0, 74.0] | 26.0 [18.0, 35.0] |  |
| Race/Ethnicity |  |  |  |
| Hispanic | 34 (10.3%) | 36 (10.9%) | 0.841 |
| Non-Hispanic White | 115 (34.7%) | 111 (33.7%) |  |
| Non-Hispanic Black | 172 (52.0%) | 168 (51.1%) |  |
| Non-Hispanic Other | 10 (3.0%) | 14 (4.3%) |  |
| Educational years |  |  |  |
| Mean (SD) | 15.4 (2.60) | 14.8 (2.39) | 0.001 |
| Median [Min, Max] | 15.0 [7.00, 21.0] | 15.0 [1.00, 21.0] |  |
| Marital Status |  |  |  |
| Married | 105 (31.7%) | 33 (10.0%) | <0.001 |
| Separated | 20 (6.0%) | 9 (2.7%) |  |
| Divorced | 72 (21.8%) | 10 (3.0%) |  |
| Widowed | 11 (3.3%) | 2 (0.6%) |  |
| Never married | 120 (36.3%) | 275 (83.6%) |  |
| Missing | 3 (0.9%) | 0 (0%) |  |
| ISS |  |  |  |
| Mean (SD) | 2.28 (1.77) | 2.34 (1.71) | 0.645 |
| Median [Min, Max] | 2.00 [1.00, 14.0] | 2.00 [1.00, 13.0] |  |
| SDS |  |  |  |
| Mean (SD) | 5.97 (8.12) | 6.43 (8.06) | 0.47 |
| Median [Min, Max] | 1.00 [0, 30.0] | 3.00 [0, 30.0] |  |
| Missing | 11 (3.3%) | 0 (0%) |  |
| ADI |  |  |  |
| Mean (SD) | 64.0 (28.9) | 60.6 (31.0) | 0.14 |
| Median [Min, Max] | 67.0 [-7.00, 100] | 64.0 [-8.00, 100] |  |
| Missing | 2 (0.6%) | 2 (0.6%) |  |
| LTS |  |  |  |
| Mean (SD) | 10.1 (9.82) | 9.90 (9.09) | 0.769 |
| Median [Min, Max] | 8.00 [0, 63.0] | 7.00 [0, 48.0] |  |
| Missing | 16 (4.8%) | 29 (8.8%) |  |
| CTQSF |  |  |  |
| Mean (SD) | 8.94 (9.96) | 9.35 (9.41) | 0.603 |
| Median [Min, Max] | 5.00 [0, 44.0] | 7.00 [0, 42.0] |  |
| Missing | 29 (8.8%) | 27 (8.2%) |  |
| PhenoAge Residuals |  |  |  |
| Mean (SD) | -0.0809 (5.70) | -0.0733 (5.92) | 0.987 |
| Median [Min, Max] | -0.0743 [-14.7, 14.4] | -0.0728 [-16.1, 13.6] |  |
| GrimAge Residuals |  |  |  |
| Mean (SD) | 0.175 (5.10) | -0.208 (4.59) | 0.311 |
| Median [Min, Max] | -0.687 [-10.7, 13.9] | -0.651 [-9.44, 12.2] |  |
| DunedinPACE Residuals |  |  |  |
| Mean (SD) | 0.00394 (0.130) | -0.00829 (0.126) | 0.221 |
| Median [Min, Max] | 0.00152 [-0.322, 0.366] | -0.0187 [-0.287, 0.401] |  |
| MDD after six months | 75 (22.7%) | 85 (25.8%) | 0.389 |

Younger participants were 35 years old or younger, older participants were older than 35 years. ADI: Area Deprivation Index, CTQSF: Childhood Trauma Questionnaire Short Form; ISS: Injury Severity Score, LTS: Lifetime Trauma Score, MDD: probable Major Depressive Disorder (assessed six months after trauma), SDS: Sheehan Disability Scale.
